# Development of a Neoadjuvant Treatment Pathway to Standardize Pancreatic Cancer Care and Improve Outcomes Across a Large Diverse Health System

**DOI:** 10.1101/2023.09.19.23295749

**Authors:** Ruwan Parakrama, Baho U. Sidiqi, Lyudmyla Demyan, Oliver Standring, Dylan J. Cooper, Shamsher Pasha, Danielle M Pinto, Tiffany Zavadsky, Xianghui Zou, Sunita Patruni, Adrianna Kapusta, Jason Nosrati, Leila T. Tchelebi, Matthew J. Weiss, Joseph M. Herman, Daniel A. King

## Abstract

**Background:** Management of localized pancreatic cancer is variable. We describe the development of a neoadjuvant therapy pathway (NATP) to standardize care across a large healthcare system.

**Methods:** We conducted an IRB-approved retrospective analysis of NATP patients between June 2019 and March 2022. The primary endpoint was NATP completion, and secondary endpoints included overall survival (OS) and quality measures.

**Results:** Fifty-nine patients began NATP, median age 70, locally advanced 44.1%. Median time on NATP was 6.1 months. The initial chemotherapy was FOLFIRINOX (64.2%) and gemcitabine/nab-paclitaxel (GnP; (35.6%)) followed by radiation in 32 (54.2%) patients. Forty-four (74.6%) completed the NATP and 30 (50.8%) underwent surgical exploration with 86.7% undergoing successful resection (61.5% R0, 23.1% R1) while 14 remained unresectable. NATP completion was associated with increased likelihood of resection (p<0.001). At median follow-up of 13.4 months, median OS was 20.9 months (95% CI 13.3- 28.5) and 1- and 2-year OS was 82.5% and 49.7%. NATP completion resulted in improved OS with median OS not reached and 1- and 2-year OS of 89.7% and 59.4% (p=0.004). Median time to NATP start was 20 days after MDR and median time to surgery was 35 days. Age, ECOG, surgical stage, chemotherapy regimen and NATP completion were significant univariable predictors of OS with ECOG status remaining significant on multivariable analysis.

**Conclusion:** Our outcomes provide a baseline for future guidance in improving care across a large system. Efforts to complete NATP and improve patient ECOG may result in more patients undergoing surgery and improve survival.

## Introduction

National guidelines for pre-operative decision making for patients with pancreatic ductal adenocarcinoma (PDAC) recommend multidisciplinary review at a high-volume center^1^. Neoadjuvant therapy improves survival for locally advanced^2^ and borderline resectable^3^ pancreatic cancer and has become increasingly adopted. Multidisciplinary care teams guide management and recommendations typically involve neoadjuvant treatment (NAT) with chemotherapy with or without radiation therapy (RT) prior to surgical exploration based on recent data supporting its use^2–10^. However, consensus on how to best deliver NAT has not yet been reached and practice is highly variable. Efforts to standardized neoadjuvant treatment are limited. For example, Moffitt et al developed an NATP limited to BRPC patients with a single restaging and only after completing NAT^11^. National standards for NAT management are lacking. NAT approaches vary across trials, with chemoradiation^2,3,5^, chemotherapy alone^8^, chemotherapy followed by chemoradiation^9^, or combination^10^. Additionally, the regimens used and number of chemotherapy cycles delivered before or after surgery has not been standardized. Treatment heterogeneity has been shown to affect outcomes in other disease sites which has resulted in the standardization of neoadjuvant approaches^12,13^ to improve the quality of care for those diseases. The management of patients with pancreatic cancer would also likely benefit from standardization of neoadjuvant approaches.

Recently, a number of high-volume centers, including Northwell Health, established a partnership with the goal of setting quality metrics to improve care^14^. Our group saw an opportunity to establish a set of standards for the management of PDAC patients with disease amenable to resection, under a standardized process we call a neoadjuvant therapy pathway (NATP). Our NATP relies on regular multidisciplinary team reviews to guide neoadjuvant therapy management. To our knowledge, a blueprint for such a pathway has not been published. In March 2019, we developed and implemented a NATP across 23 hospitals, in one of New York’s largest and most diverse health care systems. Herein we discuss the rationale behind the development of our NATP and the initial results for all patients enrolled in the first 3 years.

## Methods⍰

### 2.1 Designing the Pathway

To codify the elements of our NATP, we first assessed the practice patterns of quality variables from the literature, including large phase 2 and phase 3 trials. We compared results to practice patterns amongst oncologists within our group, and then gathered a consensus value – for example, time from biopsy to start of chemotherapy in one month, to derive each standard in our element set. We created a workflow diagram (Figure 1) which was made available at NATP meetings and shared across our health system.

**Figure 1:**
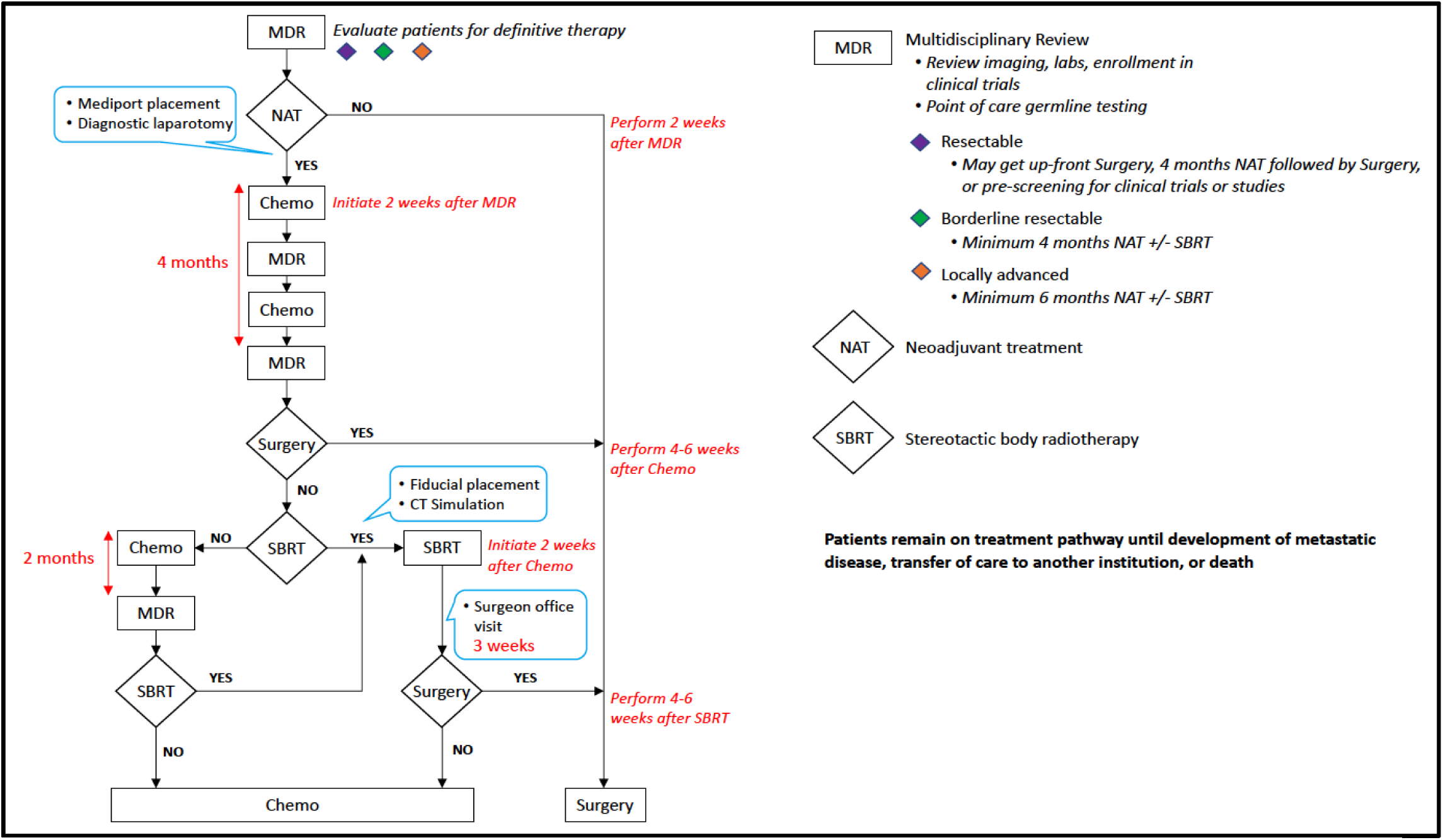
NATP decision tree highlighting the MDR prior to initiation of NAT with chemotherapy prioritized for at least 4 months before discussion for local therapies followed by further maintenance chemotherapy.

The NATP utilizes a series of coordinated MDR meetings, during which results of imaging, bloodwork, pathology, and clinical history are reviewed by an interdisciplinary team that consists of the following: coordinator, radiologist, pathologist, surgeon, medical oncologist, radiation oncologist, social worker and dietician. Patients and their family members are also encouraged to attend. Visits are arranged pre-treatment and scheduled for two and four months after the initiation of chemotherapy with imaging done prior to MDR reviews of patients. After the initial chemotherapy period, the multidisciplinary team discusses the utility and appropriateness of radiation prior to potential surgical resection. Patient eligibility for clinical trial recruitment and involvement in research studies are also discussed at each MDR review.

### 2.2 Patients

This is an institutional review board (IRB)-approved retrospective analysis of patients eligible for NATP between March 2019 and March 2022 (IRB 20-1285). The NATP tumor board consists of surgical oncologists, medical oncologists, radiation oncologists, a radiologist, a pathologist, a social worker, a palliative care expert, a dietician, a research coordinator, a biospecimen coordinator, and trainees are invited to attend. The group meets weekly for 90 minutes. On the same day as the board, patients are arranged clinic visits with a medical oncologist, and if localized disease, a surgical and radiation oncologist. Patients are also arranged appointments with a social worker and dietician. Patients are considered eligible for the NATP if, on multidisciplinary review, they have localized disease, histologically confirmed pancreatic cancer, and considered to have sufficient performance status to tolerate neoadjuvant chemotherapy. After enrollment, patients are tracked through the care process and are represented at future MDRs to ensure continued multidisciplinary discussion prior to resection. We used the National Comprehensive Cancer Network (NCCN) Pancreatic Cancer guidelines^4^ to define RPC, BRPC and LAPC patients.

### 2.3 Treatment and Clinical Pathway

Chemotherapy consisted primarily of FOLFIRINOX or GnP (gemcitabine/nab-paclitaxel). FOLFIRINOX was administered using 5-fluorouracil (5-FU); 2400 mg/m^2^), irinotecan (150 mg/m^2^) and oxaliplatin (85 mg/m^2^) every 2 weeks. GnP was administered using gemcitabine (1000 mg/m^2^) and nab-paclitaxel (125 mg/m^2^) on day 1, 8 and 15 of a one-month cycle, or on day 1 and day 15 of a monthly cycle based on tolerance and provider discretion. The choice of regimen and dose adjustments were per clinician discretion. Stereotactic body radiation therapy (SBRT) dose and fractionation options included 45-50 Gy in 5 fractions or 39-45 Gy in 3 fractions (with priority given for safety of organs at risk over coverage of tumor). Intensity modulated radiation therapy (IMRT) was used when SBRT could not be safely delivered. The surgical approach included pancreatoduodenectomy for tumors of the head and uncinate process, and subtotal or distal pancreatectomy with splenectomy for tumors of the neck, body, or tail. The final pathology included gross tumor evaluation, histological grading, lymph node involvement, margin status and tumor response based on College of American Pathologist tumor regression grading system with 0 (complete response), 1 (near complete response), 2 (partial response) and 3 (poor to no response)^15^.

### 2.4 Statistical Analysis

The primary endpoint was *completion of NATP* defined as undergoing surgical exploration or being deemed unresectable after at least 4 months of NAT. Secondary endpoints included overall survival (from start of treatment; OS), outcomes measures (resection rate, margin status, chemotherapy adherence, imaging adherence), and quality endpoints (time on NATP, delays in starting chemotherapy, and post CT/ RT completion to surgical timing). Chemotherapy adherence was defined as receiving at least 4 months of neoadjuvant chemotherapy. Imaging adherence was defined as appropriate timed imaging after 2 and 4 months of chemotherapy for MDR re-review. Full pathway adherence was defined as meeting criteria for both chemotherapy and imaging adherence. Quality metrics were initiation of RT following chemotherapy (when indicated) within 2 weeks following chemotherapy, surgery within 4-6 weeks following chemotherapy or radiation. Continuous variables were expressed using sample medians and categorical variables were expressed as percentages. The chi square method was used to compare categorical variables. The Kaplan-Meier method was used to estimate median OS. Cox proportional hazards model was used to identify independent predictors of OS. All analysis was done using SPSS (IBM SPSS Statistics for Macintosh, Version 24.0. Armonk, NY) using a P value < .05 for statistical significance.

## Results

### 3.1 The Northwell Health NATP

The NATP was designed to support biopsy-to-surgery, timely and evidence-based treatment for PDAC patients with disease potentially amenable to resection. Its core elements are summarized in Table 1 and depicted in Figure 1. There are 11 elements that are tracked within the NATP paradigm.

**Table 1.**
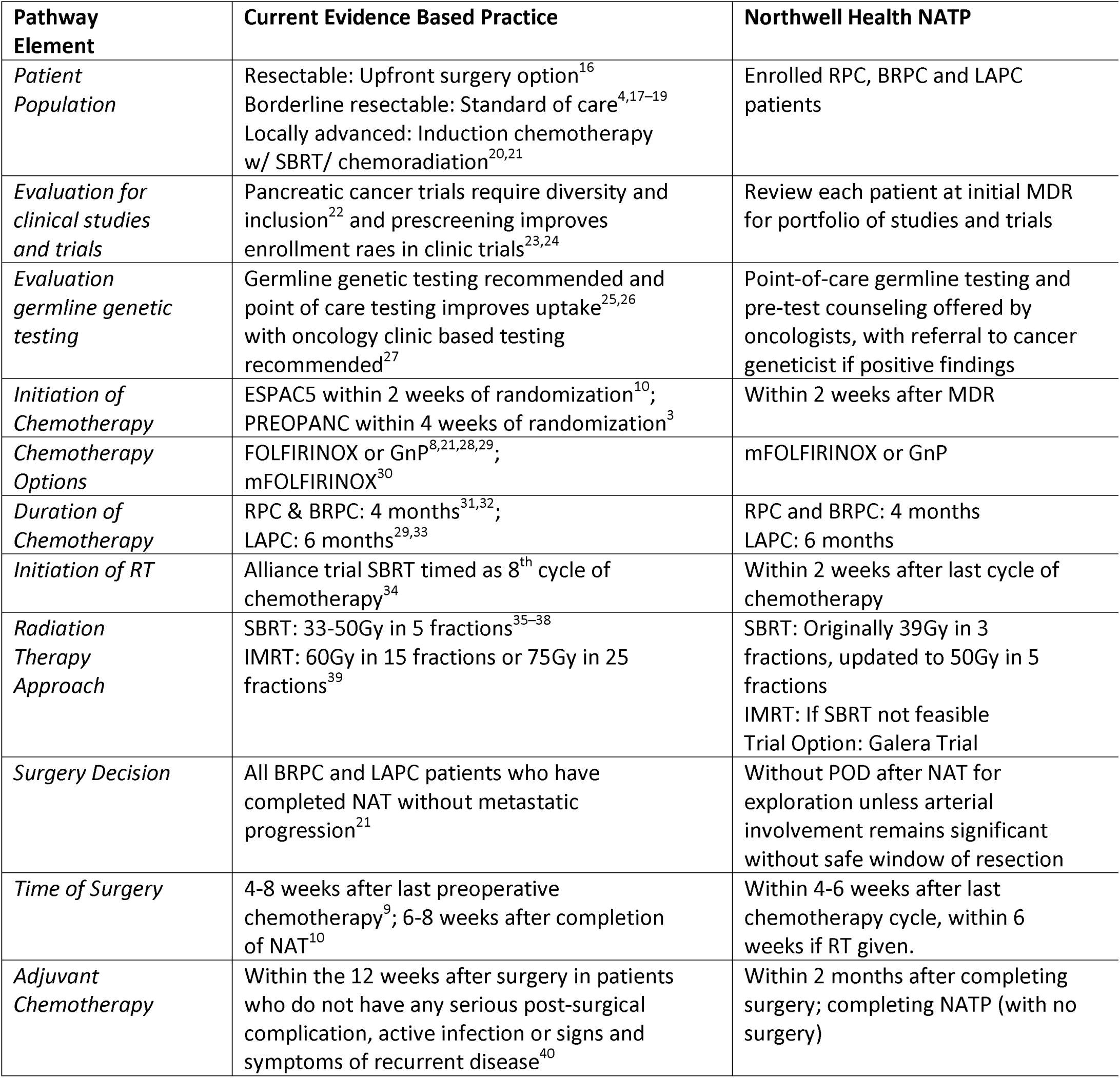
Elements of the Northwell Health NATP.

### 3.2 NATP Patient & Pathway Characteristics

There were 72 non-metastatic PDAC patients enrolled for the NATP and 59 (82%) who started treatment (Figure 2). The patient demographics of this population are represented in Table 2. Nine (15.3%) RPC, 24 (40.7%) BRPC and 26 (44.1%) LAPC patients underwent NAT. Of note, patients who self-identified as non-White represented 42% of the cohort. 15% of patients were enrolled onto clinical trials and 29% had specimens collected for biobanking.

**Figure 2:**
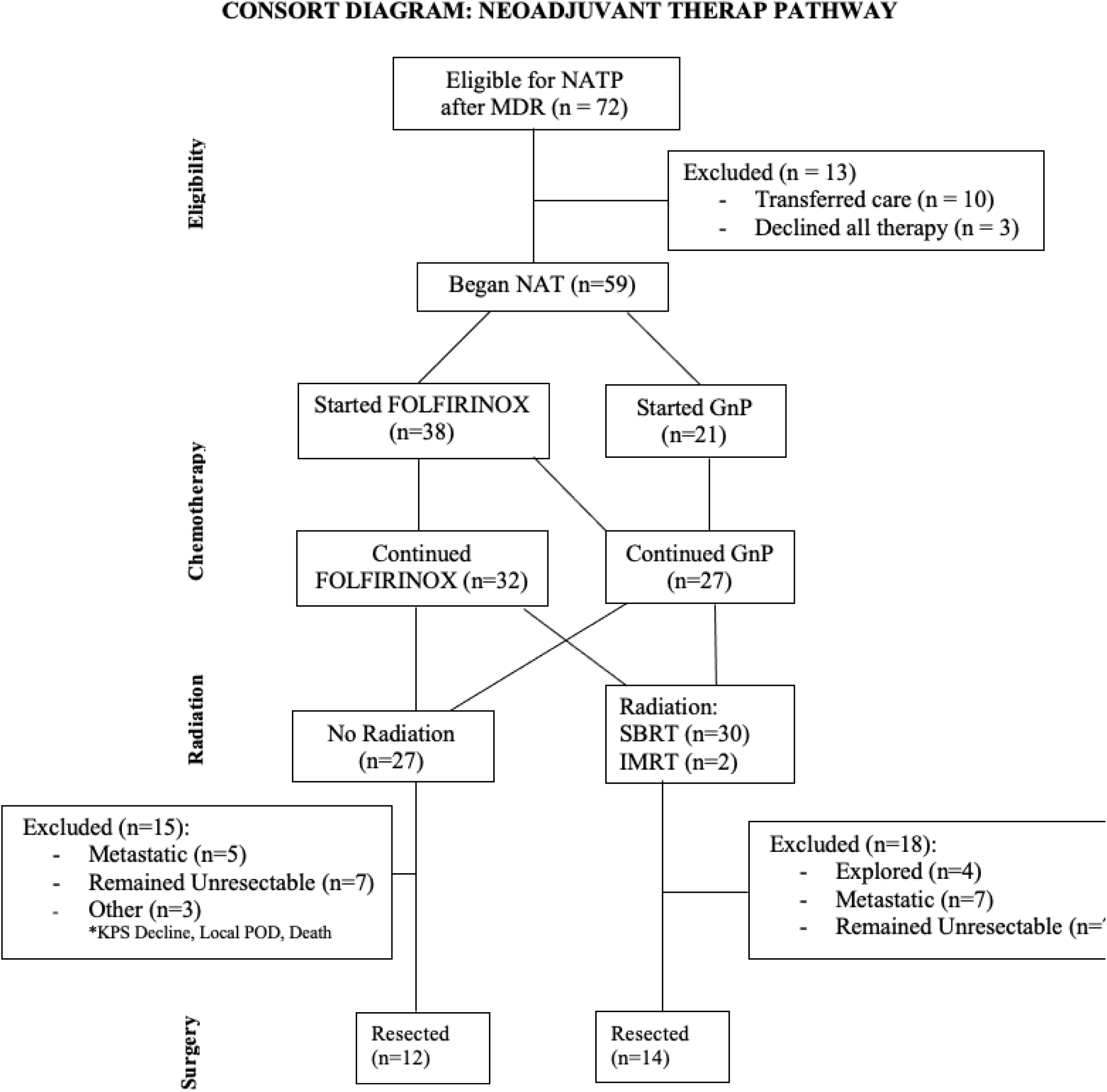
Consort diagram for NATP outcomes.

**Table 2:** Patient Demographics & NATP Metrics.

| **Characteristic** | ***n (%)*** |
| --- | --- |
| **Age (years)** |  |
| Median (mean) | 70.0 (68.5) |
| Range | 40 – 89 |
| **Gender** |  |
| Male | 27 (45.8%) |
| Female | 32 (54.2%) |
| **Race** |  |
| White | 34 (57.6%) |
| Black | 10 (16.9%) |
| Not Reported | 7 (11.9%) |
| Asian | 4 (6.8%) |
| > 1 Race | 3 (5.1%) |
| Native American | 1 (1.7%) |
| **Ethnicity** |  |
| Hispanic | 8 (12.6%) |
| Not Hispanic | 51 (86.4%) |
| **ECOG** |  |
| 0 | 23 (39.0%) |
| 1 | 27 (45.8%) |
| 2 | 9 (14.3%) |
| **Stage** |  |
| RPC | 9 (15.3%) |
| BRPC | 24 (40.7%) |
| LAPC | 26 (44.1%) |
| **MDR Presentations** |  |
| Median (mean) | 3 (2.7) |
| Range | 1 - 5 |
| **Baseline Ca 19-9** |  |
| Median (mean) | 315 (2207) |
| Range | 2 - 29865 |
| **Germline Mutation Testing** | 32 (54.2%) |
| **Clinical Trial Enrollment** | 9 (15.3%) |
| **NHBR Study Enrollment** | 23 (29.0%) |
| **Timeline Metrics** | **# (IQR)** |
| Time on NATP (months) | 6.1 (5.0 – 7.3) |
| Biopsy to Surgery (months) | 6.5 (5.7 – 8.4) |
| MDR to Surgery (months) | 6.6 (5.3 – 8.1) |
| Biopsy to CT Start (days) | 27 (21 – 41) |
| MDR to CT Start (days) | 20 (13 – 25) |
| CT Completion to RT Start (days) | 34 (22 – 42) |
| CT Alone Completion to Surgery (days) | 35 (29 – 45) |
| RT Completion to Surgery (days) | 36 (32 – 40) |
| All NAT Completion to Surgery (days) | 35 (29 – 41) |

Median time on NATP was 6.1 (IQR 5.0 –7.3) months after MDR presentation with median 20 days to initiation of chemotherapy after MDR presentation. From biopsy, median time to start of chemotherapy was 27 days with 15% of patients having biopsy post MDR review. For surgical patients, median time between biopsy to surgery was 6.5 months and from MDR presentation to surgery 6.6 months. Time to RT start after chemotherapy was 34 days, and median time to completion of all NAT to surgery was 35 days. By subgroup, last cycle of chemotherapy to surgery was 35 days and completion of RT to surgery was 36 days. All the timeline metrics are presented in Table 2 and Figure 3.

There were 38 (64.4%) patients who began chemotherapy with FOLFIRINOX, 6 (10.2%) of whom changed at the 2-month MDR to GnP due to radiographic local tumor growth or elevated Ca 19-9. A total of 307 FOLFIRINOX cycles and 167 GnP cycles were given to patients. SBRT was used in 30 (93.8%) patients with 2 patients treated with IMRT due to dose-limiting organs at risk. After completing NATP, 14 patients remained unresectable at time of reimaging. There were 30 (50.8%) patients surgically explored after NATP. Ultimately, 26 (44.1%) of the cohort underwent resection. Across all stages, completion of NAT was associated with increased likelihood of resection (p<0.001). Metastatic progression was seen in 12 (20.3%) of patients, and 3 additional patients did not complete NATP due to 1 with KPS decline, 1 with local progression and 1 death secondary to gastric outlet obstruction after 5.7 months on NATP but only 2 cycles of GnP received. The clinical treatment characteristics and pathway metrics are summarized in Table 3.

**Table 3:**
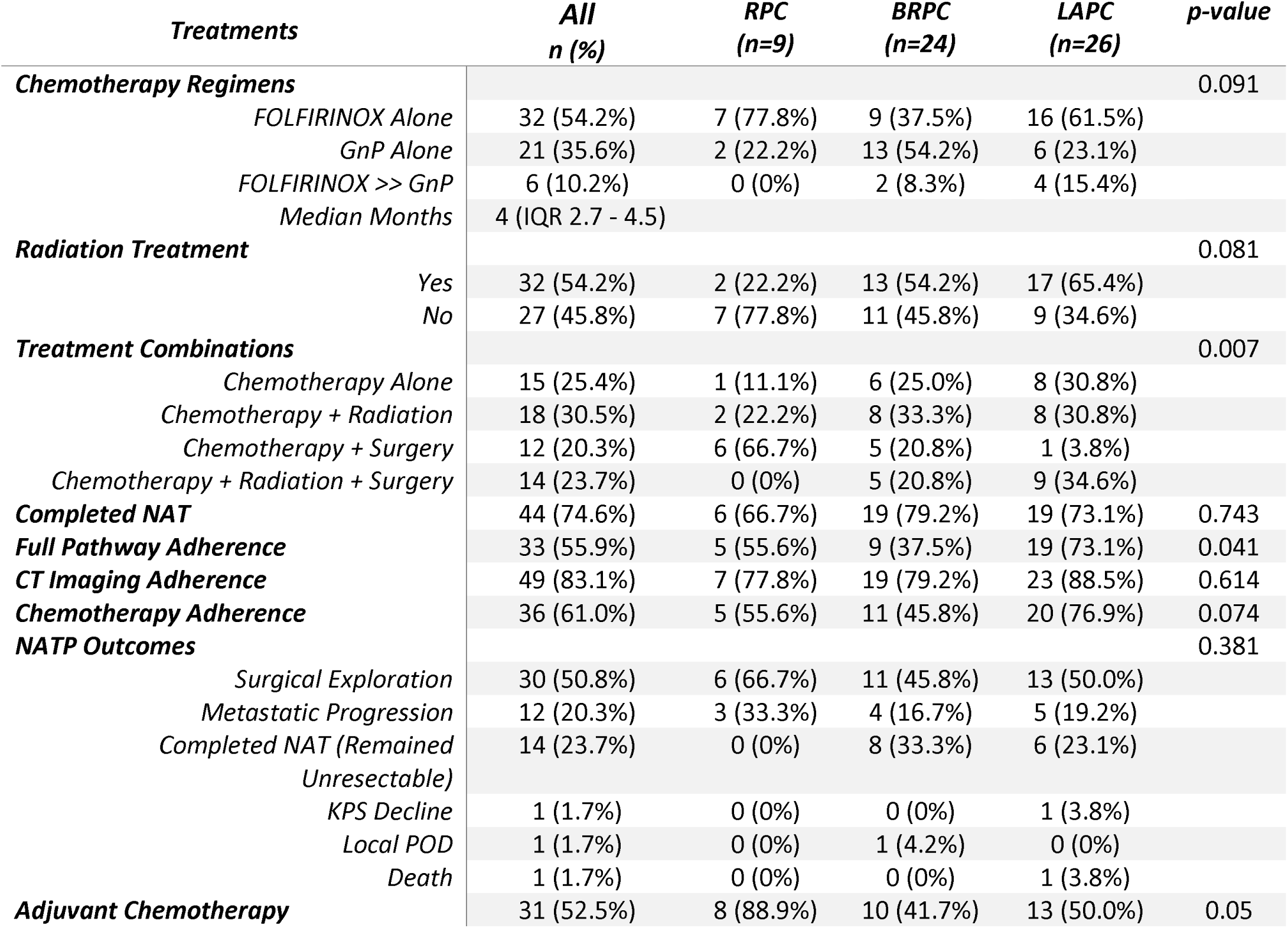
NATP Clinical Characteristics.

### 3.3 Treatment Outcomes

NATP pathologic, radiographic and clinical outcomes are summarized in Table 4. Of the 30 (50.8%) patients who underwent surgical exploration, a margin negative resection was achieved in 16 (61.5%) patients, of which 9 (56.3%) received radiation (p=0.756). Pathologic complete response (pCR) was achieved in 1 patient who underwent CT and RT and marked near complete response in 5 patients (80%) who underwent CT and RT (p=0.078). Post-surgical 30-day mortality included 1 patient death due to anastomotic leak and 1 unrelated post discharge rehospitalization due to severe COVID infection. CA 19- 9 response for all patients who were available post NATP showed a decreasing or stable CA 19-9 in 80% of patients. Additional chemotherapy was given to 31 (52.5%) patients. Individual patient level treatment course is highlighted by Figure S1 (Supplementary) swimmer plot and overall patient enrollment trends are highlighted in Figure S2 (Supplementary).

**Table 4:** NATP Treatment Response Outcomes of Patients who Underwent Resection.

| Treatment Outcomes |  | n (%) |
| --- | --- | --- |
| **Median Followup** |  | 13.4 (IQR 9.8 - 18.9) |
| **Pathology Response** |  |  |
|  | Complete Response | 1 (3.8%) |
|  | Marked Response | 5 (19.2%) |
|  | Moderate Response | 8 (30.8%) |
|  | Poor/ No Response | 11 (42.3%) |
| **Margin Status** |  |  |
|  | R0 | 16 (61.5%) |
|  | R1<1mm | 4 (15.4%) |
|  | R1 | 6 (23.1%) |
| **PNI** |  |  |
|  | Positive | 11 (42.3%) |
|  | Negative | 15 (57.7%) |
| **LVI** |  |  |
|  | Positive | 10 (38.5%) |
|  | Negative | 15 (57.7%) |
|  | Indetermined | 1 (3.8%) |
| **pT Stage** |  |  |
|  | T0 | 1 (3.8%) |
|  | T1 | 12 (46.2%) |
|  | T2 | 9 (34.6%) |
|  | T3 | 4 (15.4%) |
| **pN Stage** |  |  |
|  | N0 | 14 (53.8%) |
|  | N1 | 10 (38.5%) |
|  | N2 | 2 (7.7%) |
| **pM Stage** |  |  |
|  | Mx | 24 (92.3%) |
|  | M1 | 2 (7.7%) |
| **30 Day Post Surgery Mortality** |  | 1 (3.8%) |
| **Post Chemotherapy Ca19-9 (n=51)** |  |  |
|  | Median (mean) | 89 (889) |
|  | Range | 2 - 20811 |
| **Ca 19-9 Response (n=51)** |  |  |
|  | Decreasing | 33 (64.7%) |
|  | Stable | 9 (15.3%) |
|  | Increasing | 9 (15.3%) |
| **Additional Post NATP Chemotherapy** |  | 31 (52.5%) |

At a median follow-up of 13.4 months (IQR 9.8-8.9), median OS was 20.9 months (95% CI 13.3- 28.5). Kaplan-Meier analysis (Figure 4) showed significant differences in survival outcomes based on resection status, chemotherapy regimen and ECOG status. The 1- and 2-year OS were 82.5% and 49.7% for all patients. The Median OS after surgery was not reached (NR) vs. 16.3 months without surgery (p=0.008) with 1- and 2-year OS probability 95.7% vs. 71.6% and 64.3% vs. 31.8% respectively. The Median OS with FOLFIRINOX was not reached (NR) vs. 16.3 months (p=0.006) with 1- and 2-year OS probability 92.6% vs. 67.2% and 64.3% vs. 30.7% respectively. Median OS by ECOG status of 0, 1 and 2 was 26.1 months vs. NR vs. 12.7 months (p=0.008) with 1- and 2-year OS probability of 88.8% vs. 84.1% vs. 61.0% and 56.6% vs. 50.5% vs. 30.5% respectively. Completion of NATP also resulted in improved OS with Median OS NR vs. 15.9 months (p=0.004) with 1- and 2-year OS probability of 89.7% vs. 50.9% and 59.4% vs. 25.4%. Stage, use of radiation therapy, and pathway adherence were not significant subgroup predictors of survival. Table 5 shows OS outcomes by subgroup analysis and Figure 4 shows selected KM survival curves.

**Table 5:** Overall Survival by Subgroups.

| *Survival Outcome* | *Median OS (95% CI)* | *1 Year OS* | *2 Year OS* | *p-value* |
| --- | --- | --- | --- | --- |
| **Median OS (months)** | 20.9 (13.3 - 28.5) | 82.5% | 49.7% |  |
| **ECOG Status** |  |  |  | 0.008 |
| 0 | 26.1 (13.5 - 38.7) | 88.8% | 56.6% |  |
| 1 | NR | 84.1% | 50.5% |  |
| 2 | 12.7 (7.7- 17.7) | 61.0% | 30.5% |  |
| **Surgery** |  |  |  | 0.006 |
| Yes | NR | 95.7% | 64.3% |  |
| No | 16.3 (11.4 - 21.2) | 71.6% | 31.8% |  |
| **Chemotherapy Regimens** |  |  |  | 0.026 |
| FOLFIRINOX Alone | 26.1 (Not Specified) | 92.6% | 57.2% |  |
| GnP Alone | 16.3 (13.2 - 28.7) | 67.2% | 30.7% |  |
| **Radiation** |  |  |  | 0.793 |
| Yes | 19.1 (14.7 - 23.5) | 88.8% | 39.1% |  |
| No | NR | 73.6% | 66.9% |  |
| **Stage** |  |  |  | 0.927 |
| RPC | NR | 72.9% | 54.7% |  |
| BRPC | 26.1 (12.8 - 39.4) | 85.3% | 57.0% |  |
| LAPC | 20.9 (15.3 - 26.5) | 83.7% | 42.9% |  |
| **Completed NATP** |  |  |  | 0.004 |
| Yes | NR | 89.7% | 59.2% |  |
| No | 15.9 (10.4 - 21.4) | 60.9% | 25.4% |  |
| **Full Pathway Adherence** |  |  |  | 0.360 |
| Yes | NR | 89.7% | 48.5% |  |
| No | 26.1 (7.5 - 44.7) | 72.7% | 56.7% |  |
| **Chemotherapy Adherence** |  |  |  | 0.167 |
| Yes | NR | 90.1% | 51.2% |  |
| No | 26.1 (12.0 - 40.2) | 71.1% | 53.3% |  |
| **NATP Pathway Outcomes** |  |  |  | 0.003 |
| Surgical Exploration | NR | 96.3% | 63.8% |  |
| Metastatic Progression | 15.9 (11.1 - 20.7) | 67.3% | 18.0% |  |
| Completed NAT (Remained Unresectable) | NR | 75.0% | 56.3% |  |
| Other | 5.7 (4.6 - 6.8) | 33.3% | 33.3% |  |

**Table 6:**
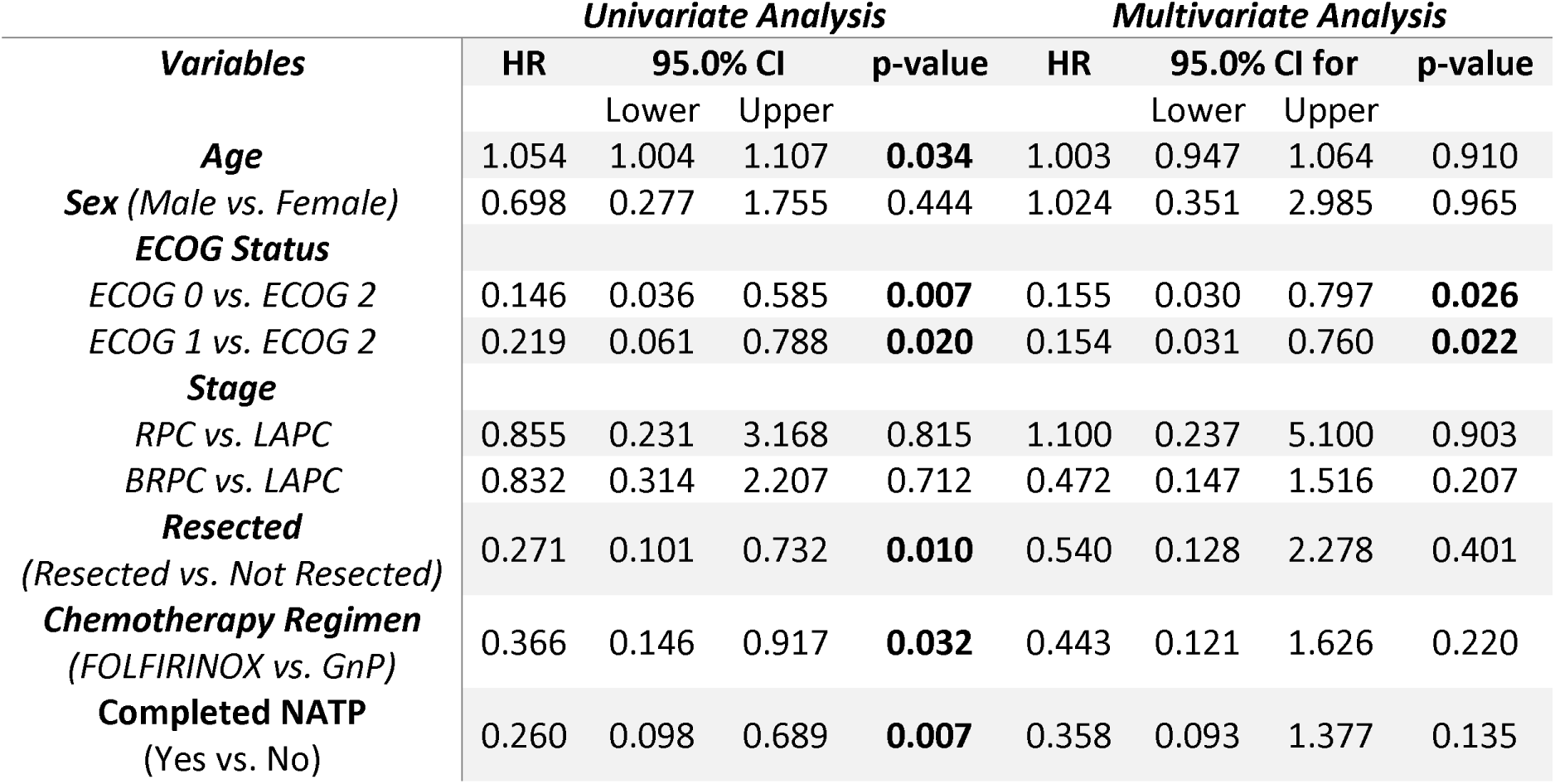
Univariate and Multivariate Analysis.

Univariate and multivariate Cox proportional hazard analysis showed age, ECOG status, surgical status, chemotherapy regimen used and completion of NATP as significant univariate predictors of OS. On MVA, only ECOG status remained a significant predictor of OS when controlling for age, gender, stage, resection status, chemotherapy use and completion of NATP. Compared to ECOG 2, both ECOG 0 (HR 0.154, 0.031-0.760; p=0.022) and ECOG 1 (HR of 0.155,0.030-0.797; p=0.026) were associated with improved survival. The full UVA and MVA analysis are summarized in Table 5.

## Discussion

Herein we present the development and implementation of a standardized NATP framework across 23 hospitals, including 6 hospitals that perform complex pancreatic surgery. We observed that patients who successfully completed all NAT were significantly more likely to undergo surgical resection and patients with the best performance status had the best long-term outcomes. The timeline metrics provide quality assurance and future guidance for quality improvement of timely care with decreasing delays.

Compliance with NCCN guidelines for the treatment of pancreatic cancer in the US is low, particularly for older people, minorities, and patients treated outside of an academic center^41^. We believe this could be improved with evidence-based standards for multimodal care, such as those proposed by the European Commissions’ Innovative Partnership for Action Against Cancer^42^. By routing new diagnoses of localized pancreatic cancer through our NATP, we provide coordinated care that benefits patients and the rest of the multidisciplinary team. We have also been able to collect data on a variety of patient outcomes – such as surgical resectability rates, response rates, as well as track outcomes. Evaluating such data will allow future work on identifying reasons for barriers in care.

There are several limitations that must be acknowledged. Our analysis is a retrospective single institution study and has associated potential biases that limit its extrapolation. Whether adherence to the NATP or better performance status are causes of good outcomes or results of chemotherapy response is not well known. The sample size is relatively small making direct comparisons difficult between subgroups and limits the ability to eliminate confounding factors when analyzing outcomes. Additionally, selection bias in selection of chemotherapy regimens, and use of radiation therapy in more advanced cases limits conclusion on best NAT approach.

However, the data presented compares well to recently published randomized trials offering support that the development of a standardized pathway offers baseline quality assurance measures for continued evaluation of our approach^2–10^. The Northwell Health Cancer Center is one of 14 centers that comprise the Canopy Cancer Collective, participating in efforts to standardize and optimize care for pancreatic cancer. The results presented here support Canopy’s efforts to promote standards for neoadjuvant therapy delivery. With respect to future directions, work is underway to better assess in real-time patients diagnosed with pancreatic cancer across our entire network, including in community oncology sites, to offer the NATP earlier at those sites. Key supportive modalities including nutrition and physical therapy are being integrated earlier into the pathway to be evaluated as part of health-related quality of life questionnaires^43^.

In conclusion, the implementation of this standardized pathway at Northwell provides the groundwork for future studies, including the continued follow-up of patients to evaluate long-term outcomes and monitoring of quality measures with focus for continued improvements. We aim to promote standardized care for pancreatic cancer across our health system at large, and other health care systems within the framework of a learning healthcare network.

## Data Availability

All data produced in the present study are available upon reasonable request to the authors.

## Notes

### Competing Interest Statement

The authors have declared no competing interest.

### Funding Statement

This study was funded in part by the 1440 Canopy Cancer Collective

### Author Declarations

IRB of Northwell Health gave ethical approval for this work

