## Supplemental Figures for "Development of a Neoadjuvant Treatment Pathway to Standardize Pancreatic Cancer Care and Improve Outcomes Across a Large Diverse Health System"

**Supplementary Figures**

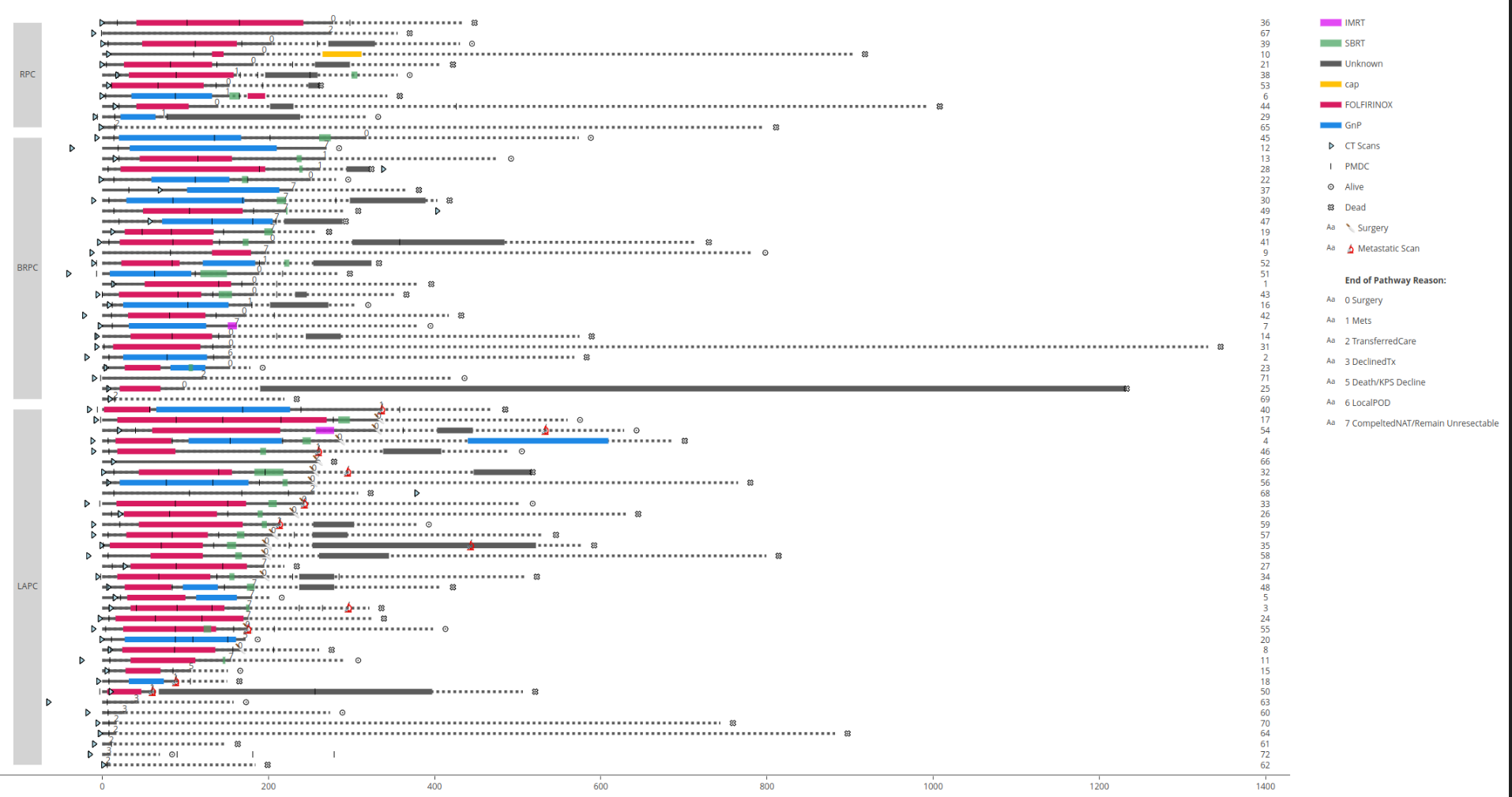

*Figure S1: Swimmer Plot highlighting individual patient treatment course*

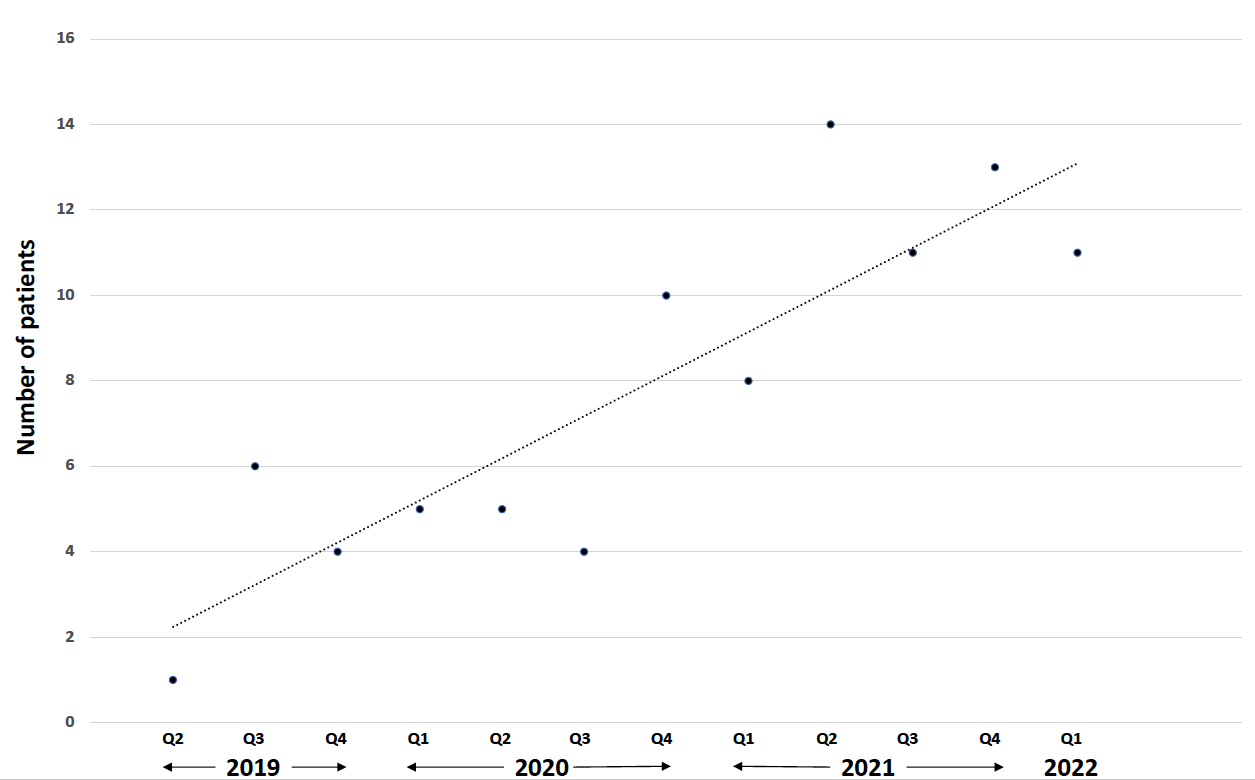

*Figure S2: Patient enrollment trends since initiation of NATP*
